# Reliability of Google Trends: Analysis of the Limits and Potential of Web Infoveillance During COVID-19 Pandemic and for Future Research

**DOI:** 10.1101/2020.12.29.20248969

**Authors:** Alessandro Rovetta

## Abstract

**Background:** Alongside the COVID-19 pandemic, government authorities around the world have had to face a growing infodemic capable of causing serious damages to public health and economy. In this context, the use of infoveillance tools has become a primary necessity.

**Objective:** The aim of this study is to test the reliability of a widely used infoveillance tool which is Google Trends. In particular, the paper focuses on the analysis of relative search volumes (RSVs) quantifying their dependence on the day they are collected.

**Methods:** RSVs of the query *coronavirus + covid* during February 1 - December 4, 2020 (period 1), and February 20 - May 18, 2020 (period 2), were collected daily by Google Trends from December 8 to 27, 2020. The survey covered Italian regions and cities, and countries and cities worldwide. The search category was set to *all categories*. Each dataset was analyzed to observe any dependencies of RSVs from the day they were gathered. To do this, by calling *i* the country, region, or city under investigation and *j* the day its *RSV* was collected, a Gaussian distribution 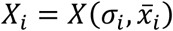 was used to represent the trend of daily variations of *x*_*ij*_ = *RSVs*_*ij*_. When a missing value was revealed (anomaly), the affected country, region or city was excluded from the analysis. When the anomalies exceeded 20% of the sample size, the whole sample was excluded from the statistical analysis. Pearson and Spearman correlations between RSVs and the number of COVID-19 cases were calculated day by day thus to highlight any variations related to the day RSVs were collected. Student t-test was used to assess the statistical significance of the differences between the average RSVs of the various countries, regions, or cities of a given dataset. Two RSVs were considered statistical confident when *t* < 1.5. A dataset was deemed unreliable if the confident data exceeded 20% (confidence threshold). The percentage increase *Δ* was used to quantify the difference between two values.

**Results:** Google Trends has been subject to an acceptable quantity of anomalies only as regards the RSVs of Italian regions (0% in both period 1 and period 2) and countries worldwide (9.7% during period 1 and 10.9% during period 2). However, the correlations between RSVs and COVID-19 cases underwent significant variations even in these two datasets (*Max* |*Δ*| = + 625% for Italian regions, and *Max* |*Δ*| = +175% for countries worldwide). Furthermore, only RSVs of countries worldwide did not exceed confidence threshold. Finally, the large amount of anomalies registered in Italian and international cities’ RSVs made these datasets unusable for any kind of statistical inference.

**Conclusions:** In the considered timespans, Google Trends has proved to be reliable only for surveys concerning RSVs of countries worldwide. Since RSVs values showed a high dependence on the day they were gathered, it is essential for future research that the authors collect queries’ data for several consecutive days and work with their RSVs averages instead of daily RSVs, trying to minimize the standard errors until an established confidence threshold is respected.

## Introduction

During the COVID-19 pandemic, fake news and inaccurate information circulated widely on the web creating severe issues to public health and economy all over the world [1]. Dr. Tedros Adhanom Ghebreyesus-director of the World Health Organization (WHO)- claimed that the battle we are fighting does not only concern the epidemic but also its infodemic [2]. Moreover, the WHO itself has launched an international campaign called “Managing the COVID-19 infodemic: Promoting healthy behaviours and mitigating the harm from misinformation and disinformation” to sensitize states to contrast the spread of misinformation [3]. To date, one of the main problems consists in conspiracy news relating to alleged vaccine damage, which can seriously compromise the international strategy for the abatement of SARS-CoV-2 [4]. Therefore, the demand for new effective and efficient infodemiological methods has never been as pressing as today. In this regard, scientists are increasingly adopting infoveillance tools to monitoring the infodemic on websites, social media, and newspapers [5]. In particular, Google Trends-an open online infoveillance tool developed by Google™ - has been widely used by the scientific community not only as for quantifying disinformation but also to make epidemiological predictions on the spread of infectious diseases, including COVID-19 [6-9]. This type of study is based on the search for statistical cross-correlations between users’ web searches related to specific diseases, such as symptoms, drugs, therapies, vaccines, number of infected people, number of deaths, etc., and the number of disease contagions and deaths officially registered after a certain timespan. However, not all that glitters is gold: indeed, Google Trends has some limitations that are often overlooked and which risk heavily biasing and distorting correlation-based analytics. Furthermore, some anomalies in the calculus of relative search volumes (RSVs) could also alter any infodemiological analysis in an unpredictable way. In this paper we will delve into the aforementioned limitations exploring their nature and searching for solutions to circumventing them, thus allowing the scientific community to continue using this precious tool through a more reliable approach.

## Methods

To assess the reliability of Google Trends (GT), relative search volumes (RSVs) of a specific query in a fixed period were downloaded on different days as to reveal any dependence on the date they were collected. In this context, “anomalies” were defined as those countries, regions or cities whose RSVs appeared only on specific days.

### Data collection

RSVs of the query *coronavirus + covid* were collected from two distinct periods: 20 February - 18 May, 2020 (period 1), and 1 February - 4 December, 2020 (period 2). Period 1, corresponding to the Italian lockdown, was chosen for GT to provide daily RSVs, while period 2 was chosen for GT to provide weekly RSVs. The survey was carried out on Italian regions and cities, and worldwide countries and cities. All RSVs of periods 1 and 2 were collected daily for a minimum of 7 days and until any anomaly was highlighted; when no anomaly was identified within 15-20 days, the investigation was considered concluded. The data-collection period ranged from 8 to 25 December, 2020. The Google Trends category search-parameter was set to *all categories*. All details are shown in Table 1.

| Geographical region | Investigation period (2020) | Subregion | Collection period (2020) | Google Trends URL [10-13] |
| --- | --- | --- | --- | --- |
| Italy | February 1 - December 4 | Regions | December 8 – 26 | URL Italy period 1 |
|  |  | Cities | December 14 – 26 |  |
|  | February 20 - May 18 | Regions | December 8 – 26 | URL Italy period 2 |
|  |  | Cities | December 14 – 26 |  |
| World | February 1 - December 4 | Regions | December 14 – 26 | URL World period 1 |
|  |  | Cities | December 16 – 26 |  |
|  | February 20 - May 18 | Regions | December 14 – 27 | URL World period 2 |
|  |  | Cities | December 16 – 27 |  |

### Statistical analysis

By calling *i* the country, region, or city under investigation and *j* the day its *RSV* was collected, a Gaussian distribution 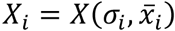, where *σ*_*i*_ is the standard deviation (also called *SD*) and 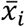 is the mean value of *RSVs*_*ij*_, was used to represent the trend of *x*_*ij*_ = *RSVs*_*ij*_. To evaluate data normality, the Shapiro-Wilk test was used [14]. The significance threshold was indicatively set at *α* = .05 [15]. Data distributions that deviated greatly from *α* were marked with an asterisk (*). The impact of daily variations of *RSVs*_*ij*_ in 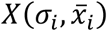 on Pearson (R) and Spearman (r) correlations with COVID-19 total cases was estimated [16]; to do this, it was enough to compute the correlations on different days and calculate their percentage increases *Δ* = (*u*_*f*_ − *u*_0_)/*u*_0_ * 100. For the adoption of these correlations, standard criteria were exploited [17]. The t-test 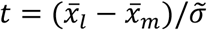 [18] was performed in order to understand if the differences between the averages of *RSVs* of the same sample (i.e. same geographical area and period) were significant. A difference between two *RSVs* was considered statistically significant when *t* < 1.5. A dataset was deemed unreliable if the confident data exceeded 20% (confidence threshold) for at least one country, region, or city. When anomalies were identified in more than 20% of cases, no investigation on the distributions was conducted.

## Results

### Italian regions’ web interest during period 1 (February – 4 December, 2020)

As shown in Figure 1, there have been strong relationships between RSVs and the dates they were collected: in fact, the regional ranking of web interest underwent several unpredictable variations even as regards the peak values *RSV* = 100.

**Figure 1.**
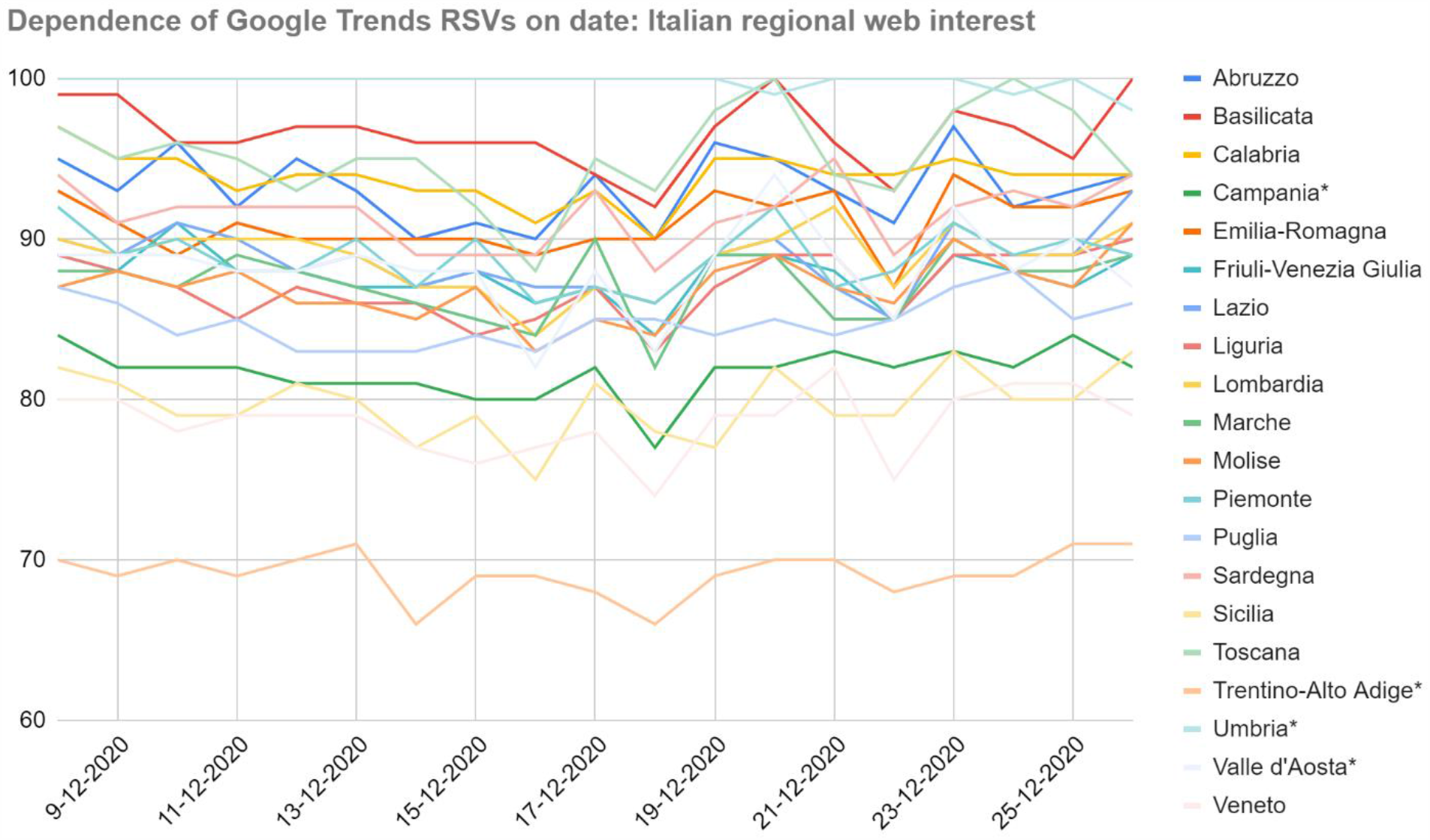
Dependence of Google Trends relative search volumes (RSVs) on collection date: Italian regions’ web interest in the query *coronavirus+covid* during period 1 (1 February – 4 December, 2020). X-axis: dates on which the RSVs were collected. Y-axis: Google Trends RSV. * = Regions that showed a non-normal trend over time.

The daily standard deviation of the sample ranged in the interval [6.6, 7.6], making all values in the central band mutually confident. Because of that, any correlations between RSVs and COVID-19 cases (or related statistics) could not be meaningful if merely based on a single-day dataset. Furthermore, even supposing no variance in daily samples, the correlation between the number of COVID-19 cases and RSVs went from *r* = .29 on December 8 to *r* = .36 on the following day (|*Δ*| = +24.1%). Considering the whole dataset, the same correlations ranged in the interval [−.23, −.42] (|*Δ*| = +82.6%). The mean value and standard error of the *X*_*i*_ distributions were 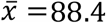 and 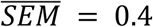 respectively, with *SEM*_*i*_ ranging in the interval [0.1, 0.7]. Therefore, the confidence threshold was exceeded (e.g. Abruzzo, 37%). However, no anomalies have been found.

### Italian regions’ web interest during the period 2 (20 February – 18 May, 2020)

As shown in Figure 2 (next page), the variance of RSVs as a function of the day they were gathered was lower than that of the previous dataset 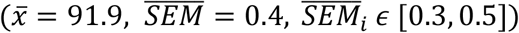. This is probably due not only to the investigated period but also to the different sampling frequency. However, there was greater variability on RSV peaks and a larger number of non-normal trends.

**Figure 2.**
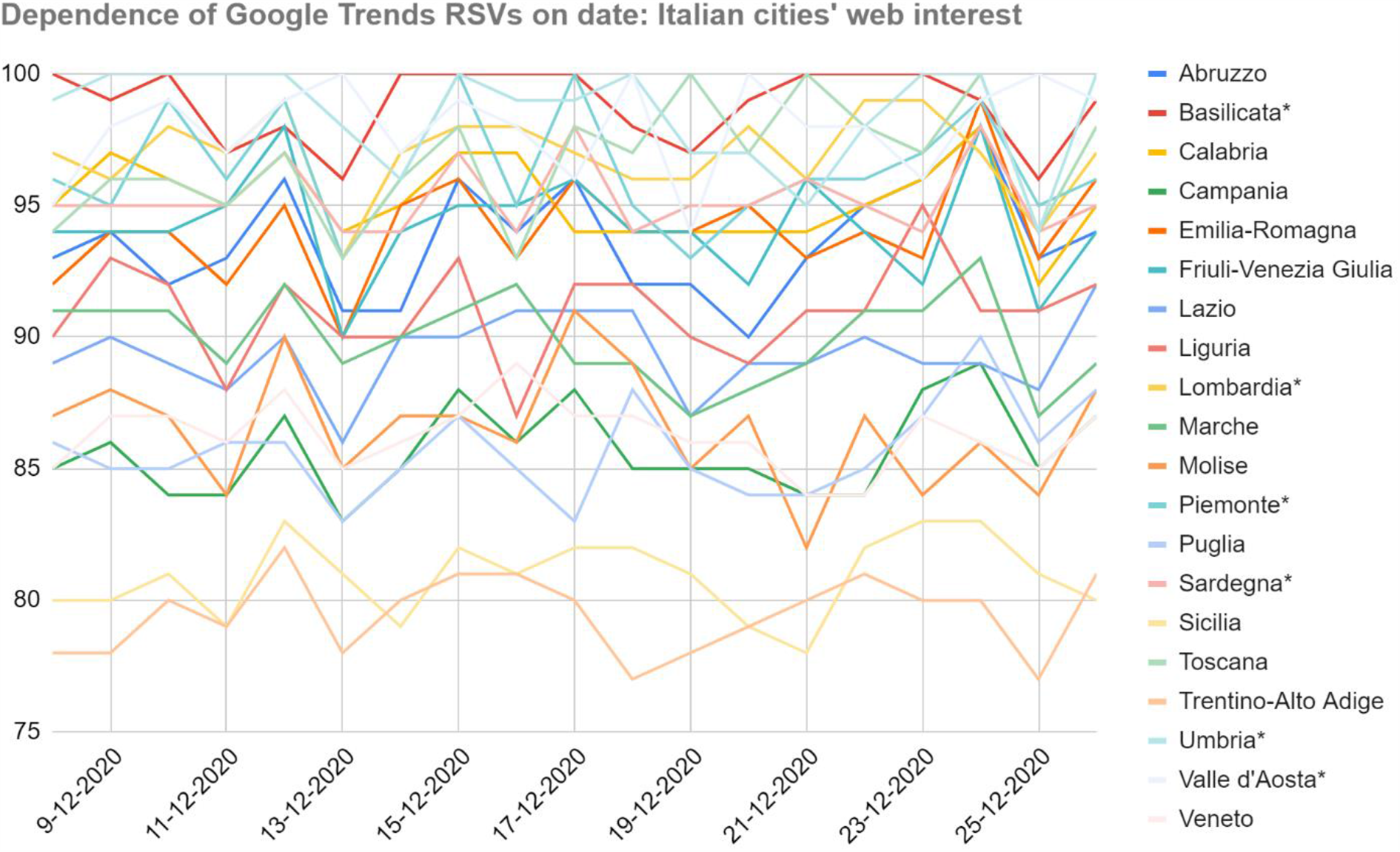
Dependence of Google Trends relative search volumes (RSVs) on collection date: Italian regions’ web interest in the query *coronavirus+covid* during period 2 (20 February – 18 May, 2020). X-axis: dates on which the RSVs were collected. Y-axis: Google Trends RSV. * = Regions that showed a non-normal trend over time.

The confidence threshold was exceeded (e.g. Abruzzo, 47%). Spearman and Pearson correlations between COVID-19 cases and daily RSVs ranged in the intervals [. 04, .29] (|*Δ*| = +625%) and [. 09, .26] (|*Δ*| = +189%) respectively. No anomalies has been found.

### Italian cities’ web interest during period 1 and period 2

As shown in Tables 2 and 3 (next two pages), significant anomalies occurred in 33.3% of Italian cities during period 1 and 45.8% during period 2. In particular, Perugia and Prato-absent respectively 7- and 10-times during period 1- recorded *RSVs* = 100 on 6 occasions. During period 2, Messina, Perugia, Pescara, Prato, and Salerno, recorded only 1 RSV out of 14 samples, while Parma recorded 2 RSVs. Therefore, any type of correlation or other statistical calculus, evaluation, or consideration on this dataset would be highly dependent on the day the data was gathered.

**Table 2.** Dependence of Google Trends relative search volumes (RSVs) on collection date: Italian cities’ web interest in the query *coronavirus+covid* during period 1 (1 February – 4 December, 2020).

| City | Weekly RSVs collected day by day from 14 to 26 December, 2020 |  |  |  |  |  |  |  |  |  |  |  |  |
| --- | --- | --- | --- | --- | --- | --- | --- | --- | --- | --- | --- | --- | --- |
| **Bari** | 94 | 98 | 94 | 94 | 97 | 95 | 94 | 96 | 94 | 95 | 91 | 91 | 94 |
| **Bologna** | 94 | 92 | 96 | 94 | 96 | 95 | 95 | 96 | 95 | 94 | 90 | 91 | 94 |
| **Brescia** | 89 | 91 | 87 | 88 | 87 | 89 | 90 | 86 | 88 | 88 | 85 | 86 | 87 |
| **Cagliari** | 97 | 98 | 100 | 96 | 97 | 95 | 94 | 98 | 96 | 96 | 97 | 91 | 98 |
| **Catania** | 88 | 89 | 86 | 82 | 86 | 86 | 89 | 85 | 85 | 88 | 84 | 83 | 87 |
| **Firenze** | 96 | 100 | 98 | 100 | 100 | 97 | 100 | 99 | 97 | 98 | 100 | 95 | 97 |
| **Genova** | 88 | 92 | 89 | 91 | 89 | 91 | 89 | 92 | 90 | 90 | 89 | 86 | 87 |
| **Milano** | 89 | 94 | 90 | 90 | 91 | 91 | 91 | 91 | 86 | 93 | 90 | 88 | 90 |
| **Modena** |  | 95 |  | 92 |  | 95 |  |  |  |  |  |  | 94 |
| **Napoli** | 89 | 93 | 89 | 88 | 91 | 90 | 91 | 87 | 90 | 94 | 88 | 84 | 89 |
| **Padova** | 88 | 90 | 88 | 87 | 91 | 88 | 87 | 84 | 87 | 89 | 87 | 84 | 87 |
| **Palermo** | 79 | 84 | 79 | 80 | 81 | 80 | 78 | 78 | 80 | 78 | 79 | 79 | 81 |
| **Parma** | 87 | 88 |  |  | 89 | 87 |  | 86 | 85 | 86 | 87 |  |  |
| **Perugia** | 100 |  |  |  |  |  | 98 | 100 | 100 |  |  | 100 | 100 |
| **Prato** |  |  |  |  |  | 100 | 100 |  |  |  |  | 95 |  |
| **Reggio Calabria** |  |  |  |  |  |  |  | 95 |  | 100 | 96 |  |  |
| **Reggio Emilia** |  |  | 88 |  | 90 |  |  |  |  |  |  |  |  |
| **Roma** | 90 | 94 | 92 | 93 | 94 | 93 | 92 | 93 | 92 | 94 | 92 | 88 | 92 |
| **Salerno** |  |  | 87 | 86 | 87 | 84 |  |  | 85 | 88 | 85 |  | 85 |
| **Taranto** |  |  |  |  |  | 100 |  |  |  |  |  |  |  |
| **Torino** | 92 | 92 | 95 | 91 | 96 | 88 | 96 | 90 | 87 | 97 | 88 | 88 | 87 |
| **Trieste** | 90 | 88 | 91 | 92 | 90 | 91 | 93 | 92 | 85 | 90 | 90 | 88 | 86 |
| **Venezia** | 82 | 85 | 83 | 81 | 80 | 83 | 81 | 82 | 79 | 84 | 80 | 79 | 80 |
| **Verona** | 83 | 85 | 86 | 81 | 86 | 86 | 86 | 84 | 82 | 84 | 82 | 79 | 82 |

**Table 3.**
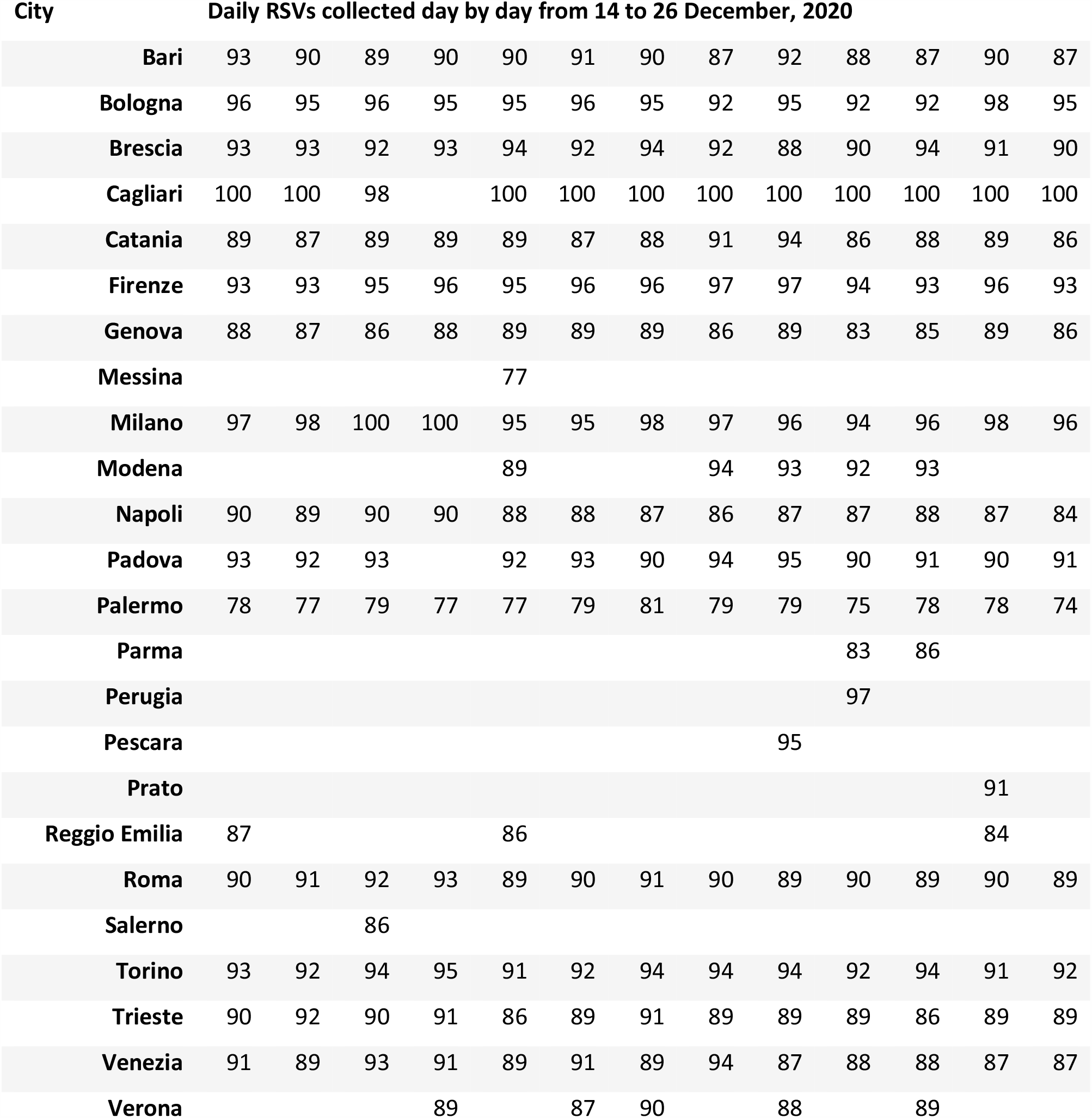
Dependence of Google Trends relative search volumes (RSVs) on collection date: Italian cities’ web interest in the query *coronavirus+covid* during period 2 (20 February – 18 May, 2020).

### Global web interest during period 1 (February – 4 December, 2020)

Google Trends reported a maximum of 62 countries’ RSVs (Supplementary Table 1). Significant anomalies occurred in 6 cases (9.7%) and the peak *RSV* = 100 was reached and maintained unchanged by Italy (*SD*_*i*_ = 0). In 64.5% of cases, data was not normally distributed. No nation exceeded the confidence threshold even if the dataset showed a high variability range if compared to that of Italy 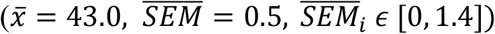. Spearman correlations with COVID-19 total cases ranged in the interval [. 04, .11] (|*Δ*| = +175%); however, it must be pointed out that the value *r* = .04 was an outlier (recorded on December 16, 2020) and a more representative interval is [. 10, .11] (|*Δ*| = +10%).

### Global web interest during the period 2 (20 February – 18 May, 2020)

Google Trends reported a maximum of 64 countries’ RSVs (Supplementary Table 2). Significant anomalies occurred in 7 cases (10.9%) and the peak *RSV* = 100 was reached and maintained unchanged by Italy (*SD*_*i*_ = 0). In 56.2% of cases, data was not normally distributed. No nation exceeded the confidence threshold even if the dataset showed a high variability range if compared to that of Italy 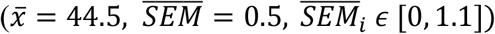. Spearman correlations with COVID-19 total cases ranged in the interval [. 04, .11] (|*Δ*| = +175%); however, it must be pointed out that the value *r* = .11 was an outlier (recorded on December 16, 2020) and a more representative interval is [. 04, .06] (|*Δ*| = +50%).

### International cities’ web interest during period 1 and period 2

As shown in Tables 4 and 5 (next two pages), significant anomalies occurred in 30.4% of international cities during period 1 and 38.1% during period 2. In particular, Bogotà, Chicago, Dubai, Houston, Hyderabad, Los Angeles, Sao Paulo, Santiago of Chile were affected by anomalies during period 1 and period 2, which also included Milan 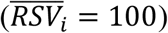 and Rome (*RSV* = 100 on December 25, 2020). Therefore, any type of correlation or other statistical calculus, evaluation, or consideration on this dataset would be highly dependent on the day the data was gathered.

**Tabel 4.**
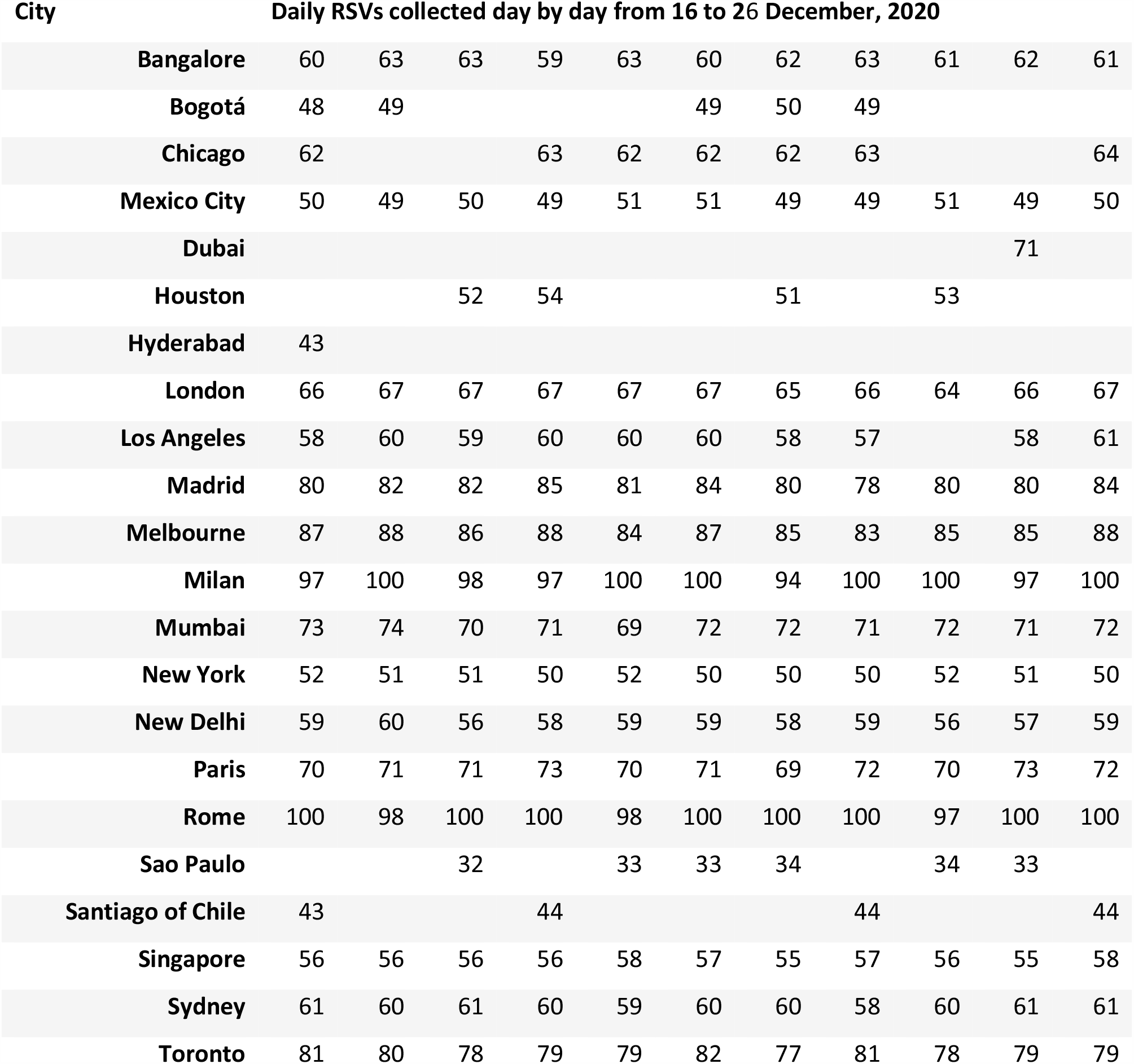
Dependence of Google Trends relative search volumes (RSVs) on collection date: international cities’ web interest in the query *coronavirus+covid* during period 1 (1 February – 4 December, 2020).

**Tabel 5.** Dependence of Google Trends relative search volumes (RSVs) on collection date: international cities’ web interest in the query *coronavirus+covid* during period 2 (20 February – 18 May, 2020).

| City | Daily RSVs collected day by day from 16 to 27 December, 2020 |  |  |  |  |  |  |  |  |  |  |  |
| --- | --- | --- | --- | --- | --- | --- | --- | --- | --- | --- | --- | --- |
| Bangalore | 67 | 67 | 65 | 67 | 65 | 66 | 66 | 66 | 70 | 68 | 65 | 65 |
| Bogotá | 50 | 52 | 49 | 51 |  | 50 |  | 51 | 52 | 53 | 50 | 48 |
| Chicago | 61 | 62 | 60 |  |  | 60 |  | 59 | 63 | 62 |  |  |
| Mexico City | 46 | 46 | 45 |  |  |  | 48 | 46 | 47 |  | 46 |  |
| Houston | 53 | 53 | 50 |  | 51 | 52 |  |  |  |  |  | 52 |
| Hyderabad |  | 49 | 48 | 49 | 48 |  |  |  |  | 50 |  |  |
| London | 64 | 64 | 64 | 63 | 64 | 65 | 66 | 64 | 67 | 65 | 63 | 62 |
| Los Angeles | 58 | 58 | 55 | 57 | 56 | 58 | 56 | 56 | 60 | 57 | 56 | 57 |
| Madrid | 83 | 85 | 85 | 83 | 82 | 85 | 86 | 84 | 84 | 86 | 83 | 87 |
| Melbourne | 60 | 61 | 60 | 58 | 58 | 60 | 59 | 58 | 62 | 64 | 58 | 60 |
| Milan | 100 | 100 | 100 | 100 | 100 | 100 | 100 | 100 | 100 |  | 100 | 100 |
| Mumbai | 78 | 77 | 75 | 76 | 76 | 76 | 76 | 78 | 78 | 80 | 77 | 77 |
| New York | 53 | 56 | 56 | 51 | 52 | 56 | 54 | 54 | 55 | 55 | 53 | 54 |
| New Delhi | 61 | 62 | 60 | 62 | 59 | 60 | 60 | 61 | 61 | 63 | 61 | 61 |
| Paris | 69 | 71 | 70 | 70 | 69 | 70 | 71 | 70 | 71 | 71 | 68 | 69 |
| Rome | 91 | 94 | 91 |  | 92 |  | 93 | 94 | 96 | 100 | 91 | 98 |
| Sao Paulo | 34 | 34 | 32 |  | 33 | 35 | 35 | 32 | 36 |  |  | 33 |
| Santiago of Chile | 45 |  | 47 |  | 46 |  | 48 |  |  |  | 46 |  |
| Singapore | 55 | 57 | 58 | 57 | 56 | 58 | 57 | 59 | 58 | 60 | 58 | 57 |
| Sydney | 55 | 56 | 56 | 56 | 54 | 55 | 55 | 56 | 57 | 59 | 55 | 57 |
| Toronto | 72 | 72 | 70 | 71 | 71 | 72 | 72 | 70 | 75 | 71 | 70 | 70 |

## Discussion

As far as the author knows, this is the first study to assess Google Trends reliability through an iterated queries analysis. In particular, this paper clearly demonstrates a strong dependence of Google Trends relative search volumes (RSVs) values on the date they are gathered. The dataset of Italian regions above all, although if not affected by anomalies, showed how the collection of the same queries’ RSVs (i.e. same category, area and period) on different days is able to substantially modify a statistical correlation between RSVs themself and an external quantity (in this case, the number of COVID-19 infections). Moreover, in all the other datasets, an even greater problem was highlighted such as the presence or absence of specific RSVs depending on the day the sample was gathered. This phenomenon has also affected cities that have reached peak values on several occasions, such as Milan and Rome in the global dataset and Perugia and Prato in the Italian dataset. Furthermore, the fact that Prato and Perugia have reached a peak of web interest in the Italian dataset but not in the international dataset shows how Google Trends RSV measurement includes only specific geographical areas according to the search item chosen by the user. Finally, RSVs of Italian regions and cities as well as RSVs of international cities showed such a daily variance that these areas were often statistically confident with each other, compromising any search for correlations or any other rank-based grouping. The most reliable dataset-i.e. a sample that showed an acceptable number of anomalies and whose data did not exceed the confidence threshold - was that of countries worldwide both during period 1 and period 2. However, even in this case there were outliers capable of destroying the correlation between RSVs and COVID-19 cases. Alongside the limitations highlighted in this paper, Cervellin et al. pointed out that web queries can be influenced by main media, further reducing the credibility of this research tool [19]. Nuti et al. have previously found that a large multitude of papers lack the information needed to make them fully reproducible [20]. Nevertheless, Google Trends has served and still serves as an excellent tool for infoveillance and infodemiology: in fact, even admitting that newspapers and newscasts can influence the trends of web queries, it provides a way to quantify the web interest in a specific topic more efficiently than any other methods historically used (e.g. population surveys) [21-24]. Moreover, it can be used as a complement to a traditional analysis [25]. In conclusion, Google Trends represents a great source of information for the entire scientific community. Nonetheless, more details should be provided by Google on how RSVs are presented to users. Finally, to ensure full reliability of a Google Trends dataset, it is essential for future research that authors collect queries’ data for several consecutive days and work with their RSVs averages instead of daily RSVs, trying to minimize the standard errors until an established confidence threshold is respected.

## Data Availability

All data can be found in the paper or in the references cited therein.

