## Supplementary tables for "Reliability of Google Trends: Analysis of the Limits and Potential of Web Infoveillance During COVID-19 Pandemic and for Future Research"

**Supplementary material**

| Country | RSVs collected from 14 to 26 December, 2020 | | | | | | | | | | | | |
| --- | --- | --- | --- | --- | --- | --- | --- | --- | --- | --- | --- | --- | --- |
| Algeria | 25 | 25 | 27 | 26 | 24 | 26 | 27 | 27 | 28 | 26 | 26 | 26 | 25 |
| Arabia Saudita | 17 | 17 | 22 | 17 | 17 | 18 | 17 | 17 | 18 | 17 | 17 | 16 | 17 |
| Argentina | 49 | 50 | 48 | 51 | 46 | 52 | 52 | 51 | 53 | 50 | 50 | 52 | 50 |
| Australia | 69 | 67 | 62 | 70 | 67 | 69 | 69 | 70 | 73 | 70 | 69 | 69 | 68 |
| Austria | 37 | 35 | 41 | 36 | 35 | 38 | 38 | 37 | 38 | 37 | 37 | 36 | 36 |
| Bangladesh | 30 | 30 | 41 | 31 | 29 | 32 | 32 | 32 | 34 | 32 | 31 | 33 | 31 |
| Belgio | 40 | 40 | 39 | 43 | 40 | 42 | 43 | 43 | 42 | 41 | 41 | 41 | 41 |
| Bolivia | 46 | 48 | 51 | 49 | 48 | 48 | 50 | 48 | 52 | 50 | 46 | 50 | 49 |
| Brasile | 32 | 33 | 34 | 34 | 32 | 34 | 34 | 35 | 34 | 32 | 34 | 33 | 34 |
| Canada | 86 | 86 | 81 | 92 | 86 | 89 | 90 | 91 | 90 | 92 | 89 | 90 | 87 |
| Cechia | 17 | 17 | 12 | 18 | 17 | 18 | 18 | 17 | 18 | 17 | 17 | 18 | 17 |
| Cile | 46 | 45 | 53 | 48 | 46 | 47 | 47 | 50 | 49 | 48 | 46 | 49 | 48 |
| Colombia | 50 | 49 | 56 | 50 | 48 | 50 | 51 | 51 | 52 | 50 | 49 | 52 | 49 |
| Costa Rica | 46 | 46 | 48 | 48 | 44 | 47 | 47 | 47 | 49 | 48 | 46 | 47 | 46 |
| Danimarca | 17 | 17 | 18 | 17 | 16 | 17 | 17 | 17 | 18 | 17 | 16 | 16 | 17 |
| Ecuador | 40 | 40 | 50 | 41 | 40 | 42 | 43 | 43 | 42 | 43 | 41 | 43 | 43 |
| Egitto |  |  | 8 |  |  |  |  |  |  |  |  |  |  |
| El Salvador | 69 | 69 | 79 | 70 | 67 | 71 | 71 | 70 | 72 | 69 | 68 | 72 | 68 |
| Emirati Arabi Uniti | 67 | 69 | 77 | 70 | 67 | 71 | 71 | 68 | 70 | 69 | 69 | 72 | 67 |
| Filippine | 30 | 30 | 34 | 30 | 30 | 30 | 31 | 32 | 32 | 30 | 31 | 30 | 30 |
| Francia | 76 | 77 | 77 | 78 | 74 | 78 | 76 | 79 | 80 | 79 | 78 | 75 | 77 |
| Germania | 30 | 30 | 37 | 31 | 30 | 32 | 31 | 31 | 32 | 31 | 30 | 32 | 31 |
| Ghana |  |  | 70 |  |  |  |  |  |  |  |  |  |  |
| Giappone | 1 | 1 | 1 | 1 | 1 | 1 | 1 | 1 | 1 | 1 | 1 | 1 | 1 |
| Grecia | 12 | 12 | 10 | 12 | 12 | 13 | 13 | 12 | 13 | 12 | 12 | 12 | 12 |
| Guatemala | 41 | 41 | 46 | 43 | 41 | 43 | 44 | 41 | 44 | 44 | 41 | 43 | 41 |
| India | 43 | 44 | 51 | 45 | 43 | 44 | 44 | 45 | 46 | 44 | 46 | 46 | 44 |
| Indonesia | 16 | 16 | 15 | 16 | 16 | 17 | 15 | 16 | 17 | 16 | 16 | 16 | 16 |
| Irlanda | 87 | 87 | 91 | 91 | 87 | 92 | 89 | 90 | 92 | 89 | 87 | 89 | 88 |
| Italia | 100 | 100 | 100 | 100 | 100 | 100 | 100 | 100 | 100 | 100 | 100 | 100 | 100 |
| Kenya | 38 | 38 | 51 | 40 | 38 | 39 | 40 | 39 | 41 | 39 | 40 | 40 | 40 |
| Malaysia | 37 | 38 | 39 | 39 | 38 | 38 | 38 | 37 | 38 | 39 | 39 | 38 | 37 |
| Marocco | 43 | 43 | 43 | 44 | 41 | 43 | 44 | 43 | 44 | 45 | 44 | 43 | 44 |
| Messico | 44 | 43 | 46 | 46 | 41 | 46 | 44 | 45 | 45 | 45 | 45 | 46 | 44 |
| Nepal | 71 | 72 | 84 | 73 | 72 | 75 | 73 | 74 | 73 | 73 | 73 | 73 | 74 |
| Nigeria | 27 | 25 | 39 | 27 | 24 | 27 | 26 | 27 | 28 | 27 | 26 | 26 | 26 |
| Norvegia |  |  | 13 |  |  |  |  |  |  |  |  |  |  |
| Nuova Zelanda | 70 | 70 | 75 | 73 | 71 | 71 | 73 | 77 | 76 | 74 | 70 | 75 | 72 |
| Paesi Bassi | 17 | 18 | 22 | 18 | 18 | 18 | 18 | 18 | 18 | 18 | 17 | 18 | 18 |
| Pakistan | 30 | 30 | 39 | 31 | 32 | 32 | 32 | 31 | 33 | 31 | 31 | 32 | 31 |
| Panamá | 49 | 49 | 60 | 50 | 50 | 51 | 51 | 50 | 52 | 51 |  | 50 | 50 |
| Perù | 61 | 61 | 68 | 63 | 60 | 63 | 64 | 63 | 64 | 64 | 62 | 63 | 62 |
| Polonia | 9 | 9 | 5 | 10 | 9 | 10 | 10 | 9 | 10 | 10 | 10 | 9 | 10 |
| Portogallo | 59 | 59 | 53 | 59 | 58 | 60 | 60 | 62 | 62 | 60 | 60 | 61 | 59 |
| Qatar | 80 | 77 | 94 | 83 | 76 | 81 | 80 | 82 | 85 | 81 | 81 | 81 | 81 |
| Regno Unito | 76 | 76 | 72 | 78 | 76 | 77 | 78 | 81 | 81 | 81 | 78 | 78 | 79 |
| Repubblica Dominicana | 32 | 30 | 39 | 32 | 30 | 32 | 32 | 32 | 34 | 32 | 31 | 32 | 31 |
| Romania | 53 | 51 | 50 | 51 | 50 | 55 | 55 | 52 | 56 | 55 | 53 | 52 | 54 |
| Russia | 2 |  | 1 |  |  |  |  |  |  |  | 2 |  |  |
| Serbia | 24 | 24 | 20 | 26 | 24 | 26 | 25 | 25 | 26 | 26 | 25 | 26 | 25 |
| Singapore | 59 | 59 | 65 | 60 | 59 | 60 | 60 | 60 | 64 | 60 | 62 | 60 | 59 |
| Slovacchia | 22 | 22 |  | 24 | 22 | 22 | 22 | 22 | 24 | 24 | 22 | 23 | 22 |
| Spagna | 81 | 80 | 91 | 82 | 80 | 82 | 82 | 82 | 84 | 83 | 82 | 81 | 82 |
| Stati Uniti | 60 | 61 | 60 | 64 | 61 | 64 | 64 | 63 | 64 | 67 | 62 | 64 | 62 |
| Sudafrica | 62 | 61 | 74 | 63 | 61 | 64 | 61 | 63 | 65 | 64 | 64 | 67 | 63 |
| Svezia | 19 | 19 | 18 | 20 | 18 | 19 | 19 | 20 | 21 | 20 | 20 | 20 | 18 |
| Svizzera | 56 | 55 | 58 | 58 | 54 | 57 | 57 | 58 | 61 | 59 | 56 | 56 | 58 |
| Thailandia | 6 | 6 | 8 | 6 | 6 | 6 | 6 | 6 | 6 | 6 | 6 | 6 | 6 |
| Turchia | 7 | 7 | 5 | 7 | 7 | 7 | 7 | 6 | 8 | 7 | 7 | 7 | 7 |
| Uruguay | 46 | 48 | 51 | 46 | 46 | 47 | 46 | 47 | 49 | 48 | 46 | 47 | 48 |
| Venezuela | 37 | 38 | 43 | 37 | 35 | 38 | 39 | 39 | 38 | 37 | 39 | 38 | 36 |
| Vietnam | 11 | 9 | 12 | 10 | 9 | 10 | 10 | 10 | 10 | 10 | 10 | 10 | 10 |

Supplementary Table 6.

| Country | RSVs collected from 14 to 26 December, 2020 | | | | | | | | | | | | | |
| --- | --- | --- | --- | --- | --- | --- | --- | --- | --- | --- | --- | --- | --- | --- |
| Algeria | 27 | 27 | 27 | 27 | 27 | 26 | 26 | 26 | 27 | 26 | 25 | 27 | 26 | 25 |
| Arabia Saudita | 20 | 22 | 18 | 22 | 21 | 21 | 20 | 21 | 22 | 21 | 21 | 22 | 21 | 20 |
| Argentina | 48 | 49 | 51 | 48 | 49 | 46 | 45 | 46 | 48 | 46 | 45 | 47 | 45 | 48 |
| Australia | 62 | 60 | 71 | 65 | 63 | 62 | 62 | 64 | 62 | 60 | 60 | 62 | 60 | 62 |
| Austria | 41 | 42 | 38 | 40 | 41 | 40 | 38 | 39 | 40 | 40 | 40 | 40 | 39 | 40 |
| Bangladesh | 41 | 40 | 32 | 42 | 41 | 42 | 41 | 42 | 42 | 43 | 40 | 42 | 41 | 41 |
| Belgio | 41 | 40 | 41 | 42 | 41 | 42 | 40 | 39 | 42 | 42 | 40 | 40 | 39 | 40 |
| Bolivia | 48 | 50 | 48 | 51 | 52 | 47 | 48 | 50 | 50 | 49 | 48 | 49 | 47 | 48 |
| Brasile | 33 | 34 | 35 | 34 | 36 | 34 | 33 | 33 | 35 | 33 | 33 | 33 | 34 | 33 |
| Canada | 80 | 80 | 92 | 84 | 85 | 81 | 81 | 82 | 81 | 79 | 83 | 81 | 79 | 82 |
| Cechia | 12 | 11 | 18 | 12 | 12 | 11 | 11 | 12 | 11 | 11 | 12 | 11 | 12 | 12 |
| Cile | 53 | 52 | 49 | 53 | 54 | 52 | 51 | 51 | 51 | 52 | 50 | 52 | 50 | 53 |
| Colombia | 56 | 55 | 51 | 56 | 56 | 53 | 54 | 53 | 53 | 55 | 53 | 54 | 53 | 54 |
| Costa Rica | 48 | 47 | 49 | 48 | 47 | 47 | 47 | 48 | 48 | 49 | 45 | 45 | 46 | 48 |
| Danimarca | 19 | 18 | 18 | 19 | 20 | 18 | 18 | 19 | 18 | 18 | 18 | 18 | 19 | 19 |
| Ecuador | 48 | 50 | 42 | 51 | 50 | 49 | 47 | 50 | 48 | 50 | 48 | 49 | 47 | 50 |
| Egitto | 8 | 8 |  | 7 | 7 | 7 | 6 | 7 | 7 | 7 | 7 | 8 | 8 | 8 |
| El Salvador | 80 | 81 | 72 | 81 | 81 | 81 | 76 | 78 | 79 | 78 | 80 | 81 | 75 | 80 |
| Emirati Arabi Uniti | 75 | 77 | 72 | 77 | 76 | 75 | 73 | 75 | 77 | 78 | 74 | 72 | 72 | 75 |
| Filippine | 32 | 34 | 31 | 33 | 34 | 31 | 31 | 32 | 33 | 31 | 33 | 33 | 32 | 32 |
| Francia | 75 | 75 | 79 | 80 | 80 | 75 | 73 | 76 | 75 | 75 | 74 | 77 | 73 | 75 |
| Germania | 38 | 37 | 32 | 37 | 38 | 37 | 36 | 37 | 37 | 36 | 36 | 37 | 36 | 37 |
| Ghana | 70 | 70 |  | 71 | 74 | 69 | 68 | 71 | 70 | 69 | 69 | 69 | 67 | 72 |
| Giappone | 1 | 1 | 1 | 1 | 1 | 1 | 1 | 1 | 1 | 1 | 1 | 1 | 1 | 1 |
| Grecia | 11 | 11 | 12 | 10 | 10 | 10 | 9 | 10 | 11 | 10 | 10 | 10 | 10 | 11 |
| Guatemala | 46 | 44 | 44 | 46 | 49 | 46 | 47 | 46 | 48 | 44 | 45 | 45 | 43 | 46 |
| India | 51 | 52 | 46 | 53 | 52 | 50 | 48 | 51 | 50 | 50 | 48 | 50 | 49 | 50 |
| Indonesia | 14 | 14 | 16 | 15 | 16 | 14 | 13 | 16 | 14 | 14 | 15 | 15 | 15 | 14 |
| Irlanda | 85 | 85 | 92 | 84 | 89 | 84 | 83 | 82 | 87 | 84 | 83 | 86 | 82 | 85 |
| Italia | 100 | 100 | 100 | 100 | 100 | 100 | 100 | 100 | 100 | 100 | 100 | 100 | 100 | 100 |
| Kenya | 51 | 52 | 40 | 54 | 52 | 50 | 50 | 51 | 53 | 50 | 51 | 50 | 52 | 51 |
| Malaysia | 38 | 39 | 38 | 37 | 38 | 37 | 36 | 37 | 37 | 36 | 36 | 37 | 35 | 37 |
| Marocco | 45 | 45 | 44 | 45 | 47 | 43 | 41 | 44 | 44 | 43 | 45 | 44 | 43 | 43 |
| Messico | 46 | 45 | 45 | 46 | 47 | 44 | 43 | 48 | 46 | 47 | 45 | 44 | 45 | 46 |
| Nepal | 80 | 81 | 74 | 84 | 81 | 81 | 79 | 82 | 83 | 81 | 80 | 81 | 80 | 80 |
| Nigeria | 40 | 39 | 27 | 39 | 41 | 39 | 38 | 39 | 38 | 39 | 37 | 38 | 38 | 38 |
| Norvegia | 14 |  |  | 15 | 14 | 14 |  |  | 14 | 13 | 13 | 13 | 13 | 14 |
| Nuova Zelanda | 74 | 75 | 74 | 75 | 78 | 73 | 73 | 76 | 75 | 75 | 74 | 74 | 73 | 75 |
| Paesi Bassi | 22 | 22 | 18 | 22 | 23 | 21 | 22 | 21 | 22 | 21 | 22 | 23 | 21 | 22 |
| Pakistan | 38 | 39 | 32 | 39 | 40 | 39 | 36 | 39 | 38 | 37 | 39 | 38 | 36 | 38 |
| Panamá | 61 | 62 | 51 | 63 | 61 | 59 | 58 | 60 | 61 | 59 | 60 | 59 | 58 | 61 |
| Paraguay |  | 50 |  | 51 | 52 |  |  | 50 |  |  |  | 50 | 47 |  |
| Perù | 69 | 67 | 64 | 71 | 72 | 69 | 68 | 67 | 68 | 69 | 68 | 69 | 67 | 67 |
| Polonia | 6 | 6 | 10 | 6 | 5 | 5 | 5 | 5 | 5 | 5 | 6 | 6 | 5 | 6 |
| Portogallo | 53 | 54 | 62 | 56 | 56 | 52 | 52 | 55 | 53 | 53 | 51 | 54 | 50 | 54 |
| Qatar | 95 | 95 | 81 | 95 | 96 | 94 | 90 | 92 | 98 | 95 | 92 | 93 | 91 | 95 |
| Regno Unito | 74 | 72 | 80 | 75 | 76 | 72 | 72 | 73 | 74 | 73 | 72 | 72 | 69 | 74 |
| Repubblica Dominicana | 40 | 39 | 32 | 40 | 41 | 39 | 37 | 39 | 40 | 39 | 39 | 40 | 39 | 40 |
| Romania | 51 | 50 | 53 | 53 | 50 | 47 | 48 | 50 | 50 | 49 | 50 | 49 | 49 | 51 |
| Russia | 1 | 1 |  | 1 | 1 | 1 | 1 | 1 | 1 | 1 | 1 | 1 | 1 | 1 |
| Serbia | 20 | 21 | 24 | 21 | 20 | 20 | 19 | 21 | 20 | 20 | 19 | 20 | 19 | 20 |
| Singapore | 62 | 65 | 62 | 65 | 63 | 62 | 61 | 60 | 61 | 60 | 62 | 62 | 60 | 64 |
| Slovacchia |  |  | 23 |  |  |  |  |  |  |  |  |  |  |  |
| Spagna | 88 | 86 | 81 | 89 | 89 | 85 | 83 | 85 | 85 | 85 | 81 | 89 | 84 | 85 |
| Sri Lanka |  |  |  |  |  |  |  |  |  | 37 | 37 |  |  |  |
| Stati Uniti | 61 | 62 | 63 | 63 | 65 | 59 | 59 | 62 | 61 | 62 | 60 | 62 | 60 | 62 |
| Sudafrica | 70 | 72 | 67 | 75 | 74 | 71 | 70 | 75 | 72 | 75 | 71 | 69 | 71 | 74 |
| Svezia | 19 | 19 | 20 | 21 | 20 | 20 | 19 | 19 | 20 | 20 | 19 | 18 | 19 | 19 |
| Svizzera | 56 | 55 | 58 | 59 | 60 | 57 | 55 | 57 | 55 | 56 | 56 | 55 | 57 | 58 |
| Thailandia | 8 | 8 | 6 | 7 | 7 | 8 | 8 | 8 | 7 | 7 | 7 | 8 | 8 | 8 |
| Turchia | 4 | 4 | 7 | 6 | 5 | 5 | 5 | 5 | 5 | 5 | 4 | 5 | 5 | 4 |
| Uruguay | 50 | 50 | 48 | 51 | 50 | 49 | 48 | 50 | 50 | 49 | 48 | 50 | 47 | 50 |
| Venezuela | 41 | 42 | 38 | 43 | 43 | 42 | 41 | 42 | 42 | 43 | 42 | 42 | 41 | 43 |
| Vietnam | 11 | 11 | 10 | 12 | 10 | 11 | 11 | 10 | 11 | 11 | 10 | 11 | 10 | 11 |

Supplementary Table 7.
